# Influenza A (H3) viral aerosol shedding in nasally inoculated and naturally infected cases

**DOI:** 10.1101/2024.09.09.24313370

**Authors:** Jianyu Lai, P. Jacob Bueno de Mesquita, Filbert Hong, Tianzhou Ma, Benjamin J. Cowling, Donald K. Milton

## Abstract

Nasally inoculated influenza cases reported milder symptoms and shed lower viral RNA load in exhaled breath aerosols (EBA) than people with classic influenza-like illness including fever, in a previous study. Whether nasally inoculated influenza is representative of mild natural influenza infection, the majority of natural infections, is unknown. Here, we extend our previous analyses to include a broader range of community-acquired influenza cases. Previously, we reported on two groups: (A) volunteers intranasally inoculated with a dose of 5.5 log_10_TCID_50_ of influenza A/Wisconsin/67/2005 (H3N2) and (B) cases with cough and sore throat plus fever or a positive rapid antigen test recruited on a college campus in the same year (2013). Here we added two additional groups from a later study: (C) cases from a 2017-2019 surveillance cohort of college dormitory residents and their contacts, and (D) cases recruited from a university health center in 2019. All cases had an influenza A(H3) infection. Using a Gesundheit-II sampler, we collected 30-minute EBA samples. Community-acquired cases from the surveillance cohort (C) shed more EBA viral RNA and were more symptomatic than the nasally inoculated cases (A) but shed less viral RNA than the natural cases that were selected for symptoms (B) in 2013, but not (D) recruited in 2019. Despite sharing a similar symptomatic profile with the 2013 selected natural cases (B), the 2019 community-acquired cases (D) recruited post-infection showed a lower fine aerosol viral RNA load. Nasal inoculation of influenza virus did not reproduce EBA viral RNA shedding or symptoms observed in mild natural infection. Circulating strains of influenza A(H3) may differ, year-to-year in the extent to which symptomatic cases shed virus into fine aerosols. New models, including possibly aerosol inoculation, are needed to study viral aerosol shedding from the human respiratory tract.

**Author Summary:** In this study, we compared influenza A (H3) viral aerosol shedding in the exhaled breath of four different groups of influenza cases: (A) volunteers given the influenza virus intranasally, naturally infected (B) college community members with classic influenza-like illness including fever recruited in 2013, (C) dormitory residents undergoing active surveillance, and (D) patients from a university health center recruited in 2019. We found that mild symptomatic cases among healthy college students (C) released more viral RNA in their exhaled breath than those nasally inoculated with influenza virus (A). We also observed that more symptomatic and medically attended cases from different flu seasons (B and D), although reporting similar symptom severity, shed different levels of viral RNA in their exhaled breath. Our findings indicate that influenza viral aerosol shedding varies from season to season. Most volunteers nasally inoculated at a high virus dose did not shed detectable viral RNA in their exhaled breath or show symptoms, suggesting that nasal inoculation may not accurately mimic natural infection. Our results highlight the need for improved models to study the spread of influenza virus in aerosol forms.

## Introduction

Seasonal influenza virus infections impose a tremendous health burden (1–4). In the United States alone, Centers for Disease Control and Prevention estimated that influenza-related deaths ranged from 12,000 to 52,000 per season from 2010 to 2020, with between 140,000 and 710,000 hospitalizations per season (5). Among the influenza strains, A(H3N2) viruses often result in more severe disease burdens in terms of morbidity and mortality compared to other strains, such as A(H1N1) and B viruses (2).

Existing research demonstrates the major role of aerosol inhalation in influenza transmission (6,7). One study estimated that approximately half of influenza transmissions in households occur via inhalation of aerosols (6). Experimental influenza cases infected via intranasal inoculation of viruses typically shed lower levels of fine and coarse EBA than naturally-infected symptomatic influenza cases (7,8), and experience milder symptoms compared to those infected naturally (8,9) or via aerosol inoculation (10). Influenza A viral shedding quantity typically aligns with the progression of clinical symptoms over the course of the disease (8,11). For instance, the peaks of EBA viral shedding and symptom scores were observed on day one post symptom onset for selected natural cases and on the third day post-inoculation for experimental cases (8).

Prior research comparing viral shedding between natural and intranasally inoculated influenza cases has primarily focused on naturally infected cases presenting with acute respiratory illness, a fever or a positive rapid antigen test (8,9). This tendency to recruit symptomatic cases from the population for aerosol shedding studies led to a lack of data on those with mild or asymptomatic influenza viral infection. Additionally, there has been limited comparative research on EBA viral shedding trajectories for both natural cases unselected for symptoms and experimental nasal inoculated cases. Despite observations that experimentally infected cases shed less virus in EBA than naturally infected cases, there is a lack of overlap in symptom severity between the two groups, with the experimentally infected cases mostly presenting with mild illness or asymptomatically (8). The observed relationship between symptom severity and exhaled breath viral shedding was potentially confounded by the distinct populations under study, one being naturally infected and the other infected via nasal inoculation.

This study sought to address these limitations by broadening the scope of prior analyses. We included unselected community-acquired influenza cases from a surveillance cohort of college dormitory residents and their contacts. This group was defined as “unselected” due to being enrolled and followed up for cold or flu-like symptoms regularly, with validated incentives for consistent responses (12). We also included cases recruited post infections from a university health center, who generally presented with more symptoms. We compared these two groups with volunteers intranasally inoculated with influenza A/Wisconsin/67/2005(H3N2) as well as selected naturally infected influenza cases with cough and sore throat plus fever or a positive rapid antigen test recruited in 2013 that were reported previously (8). Our aim was to examine the EBA viral shedding in an unselected sample of community-acquired influenza cases and compare the EBA viral RNA loads and their trajectories between nasally inoculated cases and different community-acquired influenza cases.

## Results

### Characteristics of the study population

This study included a total of 143 participants infected with influenza A(H3), with 36 nasally inoculated with influenza A/Wisconsin/67/2005(H3N2) (Group A), 83 selected symptomatic cases with fever or positive antigen test recruited in 2013 (Group B), 17 unselected cases recruited from the surveillance dorm resident cohort (Group C), and 7 symptomatic cases recruited from a university health center (UHC) in 2019 (Group D) (Figure 1). Group A had the most participants over 25 years of age (mean age: 30.1 years), while the other three groups predominantly consisted of younger adults, aged 18 to 25 years. A higher proportion of females was found in Groups B (57%) and C (65%) compared to Groups A (31%) and D (29%) (Table 1 and Supplementary Table 1).

**Figure 1.**
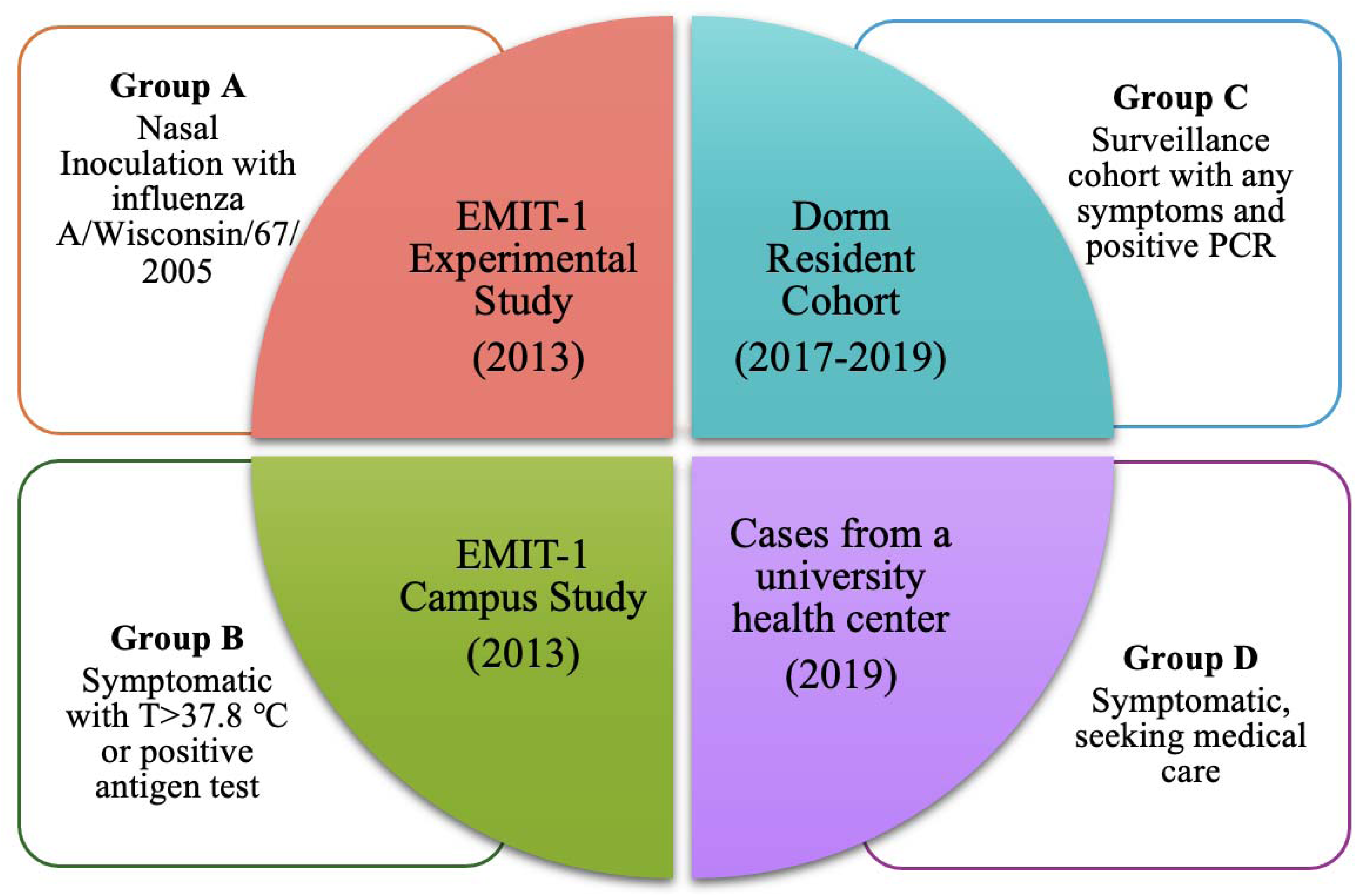
Study population

**Table 1.** Characteristics of the study population.

|  | A: Nasal<br>Inoculation<br>with<br>influenza<br>A/Wisconsin<br>/67/2005 | B:<br>Symptoma<br>tic with<br>T>37.8°C<br>or positive<br>antigen<br>test | C:<br>Surveillan<br>ce cohort<br>with any<br>symptoms<br>and<br>positive<br>PCR | D:<br>Symptomat<br>ic, seeking<br>medical<br>care | All<br>participa<br>nts |
| --- | --- | --- | --- | --- | --- |
| Number of | 36 | 83 | 17 | 7 | 143 |
| participants |  |  |  |  |  |
| Influenza<br>Season,<br>N(%) |  |  |  |  |  |
| 2012-2013 | 36 (100) | 83 (100) | 0 (0) | 0 (0) | 119 (83) |
| 2016-2017 | 0 (0) | 0 (0) | 3 (18) | 0 (0) | 3 (2) |
| 2017-2018 | 0 (0) | 0 (0) | 4 (24) | 0 (0) | 4 (3) |
| 2018-2019 | 0 (0) | 0 (0) | 10 (59) | 7 (100) | 17 (12) |
| Female,<br>N (%) | 11 (31) | 47 (57) | 11 (65) | 2 (29) | 71 (50) |
| Age,<br>mean ± SD | 30.1 ± 7.2 | 22.3 ± 7.6 | 19.3 ± 0.9 | 20 ± 1.4 | 23.8 ± 7.8 |
| Age group,<br>N(%) |  |  |  |  |  |
| <18 | 0 (0) | 2 (2) | 0 (0) | 0 (0) | 2 (1) |
| 18-25 | 11 (31) | 73 (88) | 17 (100) | 7 (100) | 108 (76) |
| >25 | 25 (69) | 8 (10) | 0 (0) | 0 (0) | 33 (23) |
| With fever ><br>37.9°C,<br>N (%)<sup>a</sup> | 1 (3) | 17 (20) | 2 (12) | 1 (14) | 21 (15) |
| Coughs per<br>30 min,<br>mean ± SD<br>(range)<sup>b</sup> | 2 ± 6<br>(0-35) | 27 ± 33<br>(0-265) | 13 ± 18<br>(0-66) | 15 ± 21<br>(0-60) | 19 ± 29<br>(0-265) |
| Temperature<br>(C),<br>mean ± SD<sup>c</sup> | 36.6 ± 0.6 | 37.3 ± 0.6 | 37.3 ± 0.8 | 37.6 ± 0.7 | 37.2 ± 0.7 |
| Median<br>upper<br>respiratory<br>symptoms<br>(IQR)<sup>d</sup> | 1<br>(0 - 3) | 7<br>(5 - 9) | 6<br>(4 - 7) | 5<br>(3.5 - 6.5) | 5<br>(3 - 8) |
| Median<br>lower<br>respiratory<br>symptoms<br>(IQR) | 0<br>(0 - 0) | 3<br>(2 - 4) | 3<br>(2 - 3) | 2<br>(2 - 4.5) | 2<br>(1 - 4) |
| Median<br>systemic<br>symptoms<br>(IQR) | 0<br>(0 - 1) | 5<br>(4 - 7) | 3<br>(1 - 6) | 6<br>(4 - 7.5) | 4<br>(1 - 6.5) |

- 126 a. Fever at the time of breath sample collection. Tympanic temperature was measured for Group A and oral temperature was measured for Groups B, C, and D. One subject in Group C did not have body temperature measured. b. Cough counts were missing for 4 observations in Group A, 3 observations in Group B, and 3 observations in Group C.
- 131 c. Temperature was missing for 1 observation in each of Groups A, B, and C. d. Upper respiratory symptoms (range 0-15): runny nose, stuffy nose, sneezing, sore throat, and earache symptom scores; lower respiratory symptoms (range 0-6): shortness of breath and cough; systemic symptoms (range 0-9): malaise, headache, and muscle/join ache.

As previously reported (8), Group A had very mild or non-existent symptoms. This group had the lowest median score for all the three symptoms – upper respiratory symptoms, lower respiratory symptoms, and systemic symptoms. They also had the lowest incidence of febrile cases (3%) and a reduced frequency of coughs at the time of sampling. Group C, the unselected community-acquired cases from the surveillance cohort, demonstrated a medium level of symptom severity across the groups. Specifically, we found a significant difference among the three community-acquired infection groups in terms of their systemic symptoms and a borderline significant difference for upper respiratory symptoms (Supplementary Table 1). Group C had significantly lower upper respiratory and systematic symptom scores than Group B. For Group B and D, the symptom distribution, as well as other characteristics reported, did not demonstrate substantial differences (Table 1, Supplementary Table 1, and Supplementary Figure 1).

### Positive detection rates and geometric means of EBA viral shedding

Group A demonstrated a significantly lower rates of positive detection for both coarse and fine EBA, in terms of the number of positive samples and positive cases, than Group B and D. Group C had a slightly higher proportion of positive cases and samples than Group A, although the differences were not significant.

There was no significant difference between Groups B and D in the proportions of positive cases and positive EBA samples. When comparing to Group C, we only found significant difference in the rates of positive cases and samples for fine EBA between Groups B and C (Table 2).

**Table 2.** Viral shedding into exhaled breath aerosols.

|  | A: Nasal Inoculation<br>with influenza<br>A/Wisconsin/67/2005 |  | B:<br>Symptomatic<br>with<br>T>37.8°C or<br>positive<br>antigen test |  | C:<br>Surveillance<br>cohort with<br>any<br>symptoms<br>and positive<br>PCR |  | D:<br>Symptomatic,<br>seeking<br>medical care |  |
| --- | --- | --- | --- | --- | --- | --- | --- | --- |
| Year | 2013 |  | 2013 |  | 2017-2019 |  | 2019 |  |
| EBA | Coarse | Fine | Coarse | Fine | Coarse | Fine | Coarse | Fine |
| Cases | 36 | 36 | 83 | 83 | 17 | 17 | 7 | 7 |
| Samples | 64 | 64 | 143 | 143 | 21 | 21 | 7 | 7 |
| No. of<br>positive<br>cases<br>(%) | 6 (17) | 11 (31) | 43<br>(52) | 71<br>(86) | 5 (29) | 8<br>(47) | 4 (57) | 6<br>(86) |
| No. of<br>positive<br>samples<br>(%) | 6 (9) | 14 (22) | 64<br>(45) | 110<br>(77) | 5 (24) | 8<br>(38) | 4 (57) | 6<br>(86) |

Figure 2 shows geometric mean viral RNA copy numbers measured in aerosols collected per 30-mimute sampling period. In this analysis without controlling for confounders, Group A had a significantly lower geometric mean viral RNA copy numbers in both fine and coarse EBA, relative to the other three groups (Figure 2). Group B presented a significantly higher geometric mean viral RNA copy numbers in fine, but not in coarse EBA, compared to Group C and D. Both Group C and D displayed intermediate levels of fine EBA viral RNA load, although D had a higher value than C.

**Figure 2.**
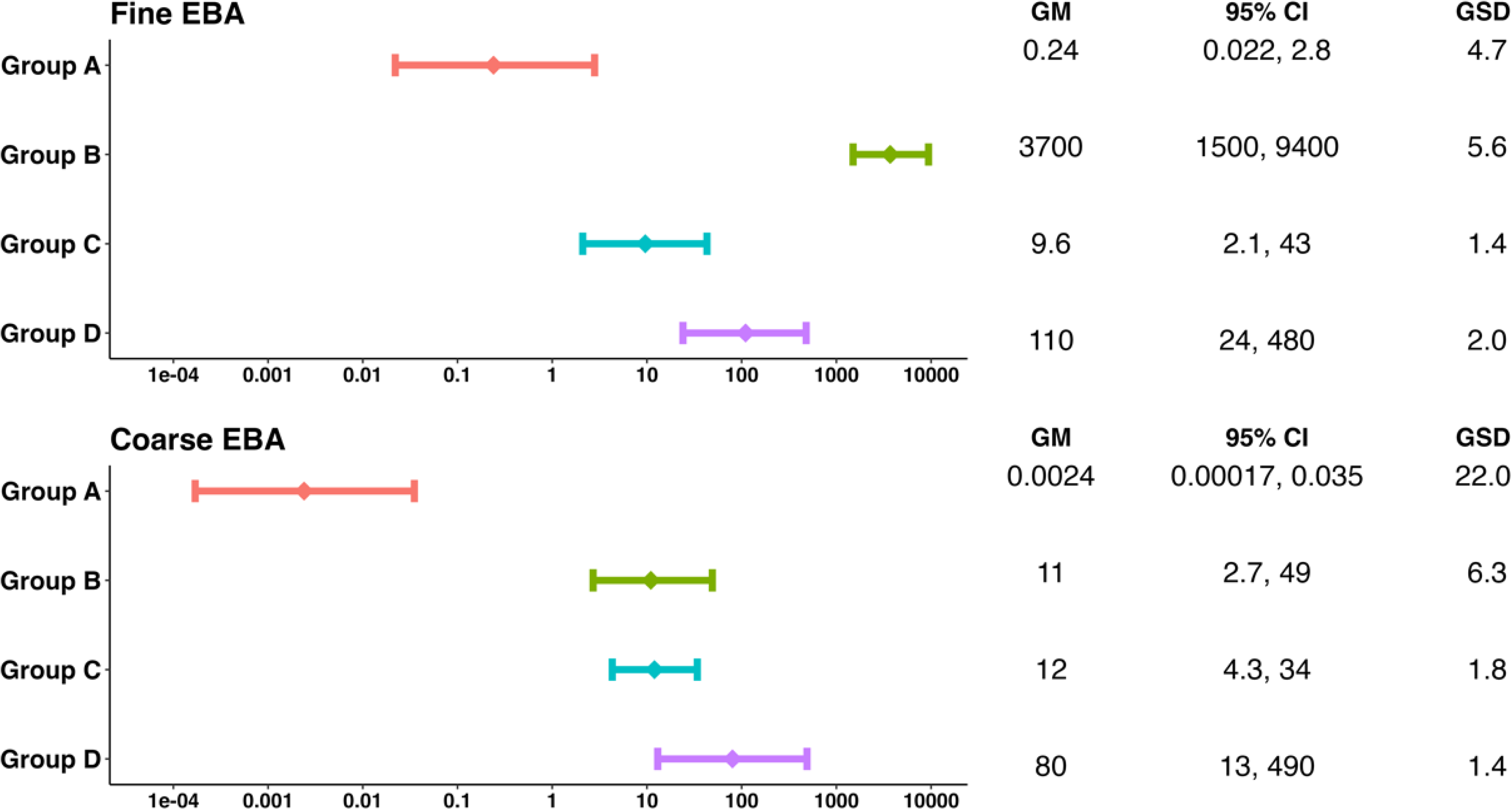
Geometric means of EBA viral shedding for the four groups. GM, geometric means; GSD, geometric standard deviation. Group A: Nasal Inoculation with influenza A/Wisconsin/67/2005; Group B: Symptomatic with T>37.8°C or positive antigen test; Group C: Surveillance cohort with any symptoms and positive PCR; Group D: Symptomatic, seeking medical care. The GM and GSD were computed for all the samples using linear mixed-effects models for censored responses (R Project package “lmec”). These models accounted for the censored outcome variable as well as nested random effects of individuals and samples within the same individuals.

### Temporal Patterns of Symptoms and EBA Viral RNA Shedding

For Groups B, C, and D, symptom scores peaked on the first day following symptom onset, with the exception of upper respiratory symptom scores in Group C, which fluctuated post-onset. For Group A, all symptom scores reached their peak on the third day post-inoculation. Based on the observed trajectories, we estimate an incubation period of two days for Group A (Figure 3).

**Figure 3.**
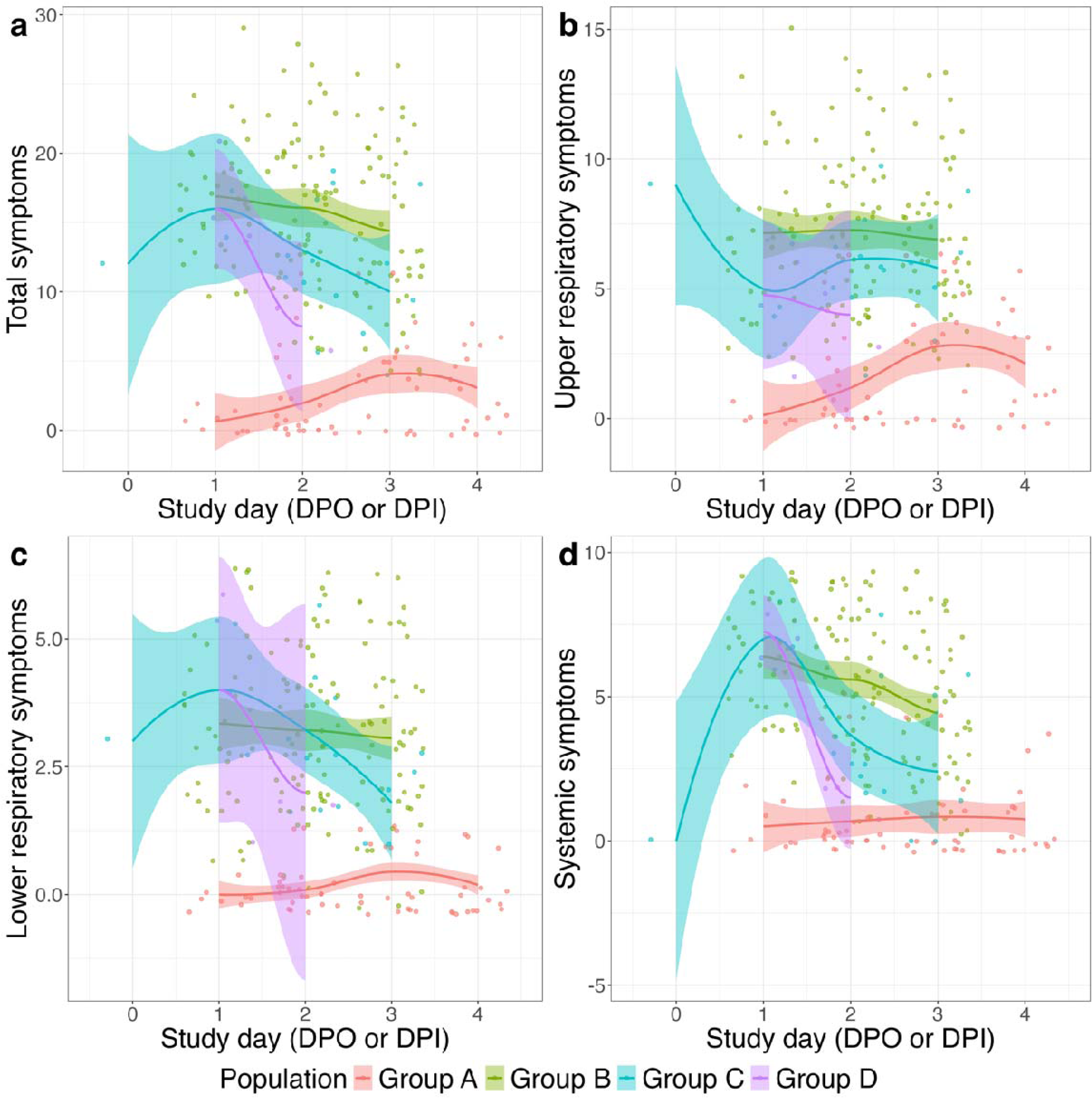
Trajectories of symptom scores over study days. Group A: Nasal Inoculation with influenza A/Wisconsin/67/2005; Group B: Symptomatic with T>37.8°C or positive antigen test; Group C: Surveillance cohort with any symptoms and positive PCR; Group D: Symptomatic, seeking medical care. Study day was defined as day post symptom onset for Group B, C, and D, and day post inoculation for Group A.

The trajectories of EBA viral RNA shedding mirrored those of the symptom scores, with the exception of Group C. For this group, the EBA shedding trajectory followed the symptom trajectory for the initial two days, but then increased on the third day post-symptom onset (Figure 4).

**Figure 4.**
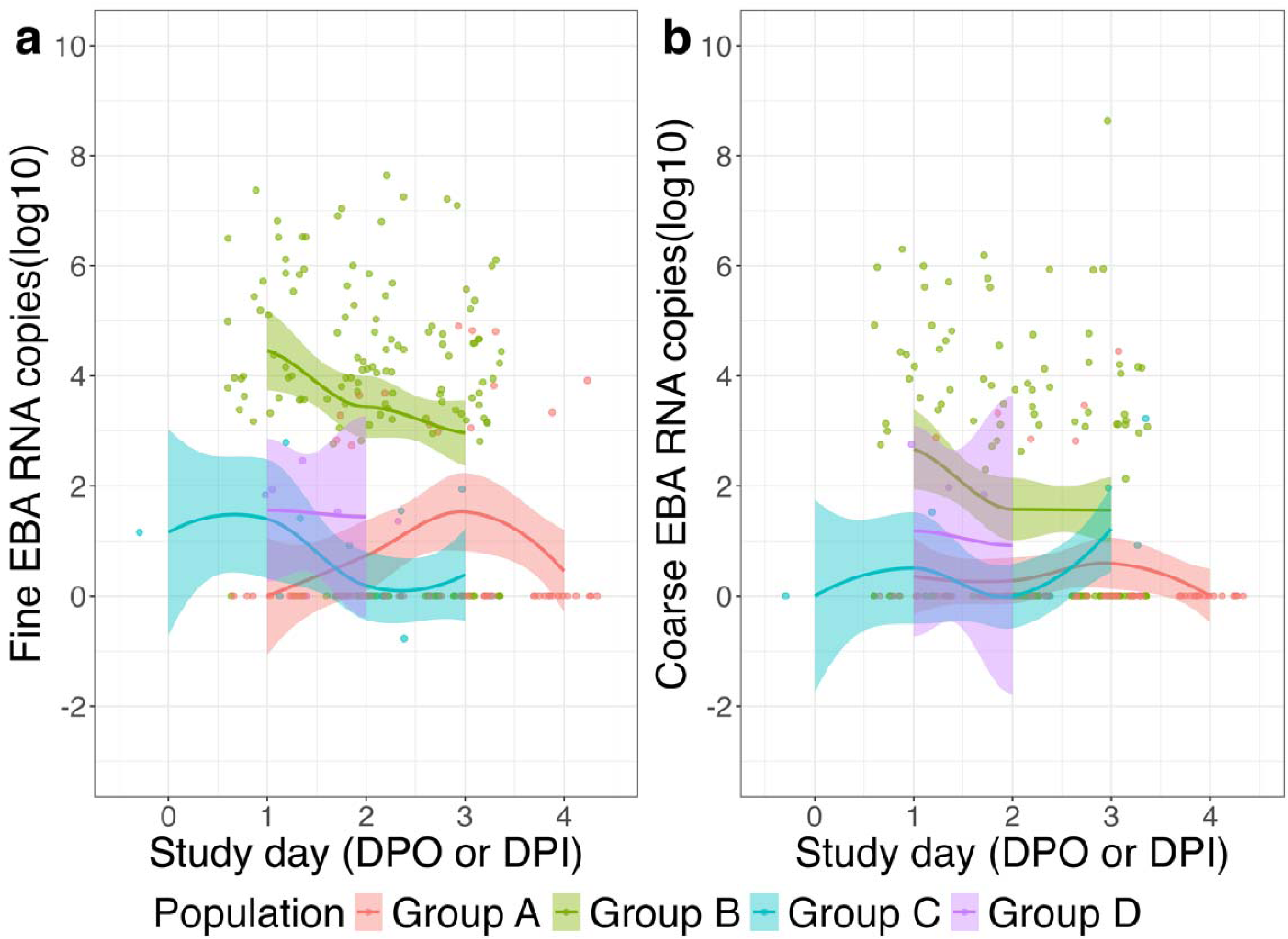
Trajectories of EBA viral shedding over study days. Group A: Nasal Inoculation with influenza A/Wisconsin/67/2005; Group B: Symptomatic with T>37.8°C or positive antigen test; Group C: Surveillance cohort with any symptoms and positive PCR; Group D: Symptomatic, seeking medical care. Study day was defined as day post symptom onset for Group B, C, and D, and day post inoculation for Group A.

### Modeling Group Effects on EBA Viral RNA Shedding

In models controlling for potential confounding variables and effect modifiers (age, sex, and study days), Group A continued to present a significantly lower viral RNA load in both aerosol fractions. The ratio of viral RNA shedding of Group A to that of Group C was the lowest, with a value of 3.3×10^−2^ (95%CI: 1.1×10^−3^, 0.99) for fine aerosol and 9.5 x 10^−4^ (95%CI: 4.4 x 10^−6^, 0.21) for coarse aerosol. Group B, on the other hand, had a significantly higher viral RNA load in both aerosols compared to Group C, with the ratio as 2.2×10^3^ (95%CI: 1.7×10^2^, 2.8×10^4^) for fine and 88 (95%CI: 1.6, 4.8×10^3^) for coarse aerosols. There was no significant difference between Group C and Group D in terms of viral RNA load in both EBA size fractions (Figure 5 and Supplementary Table 2). Contrast analysis revealed that Group B had a significantly higher viral load in fine aerosols than Group D, although no significant difference was found for coarse aerosols (Supplementary Table 3).

**Figure 5.**
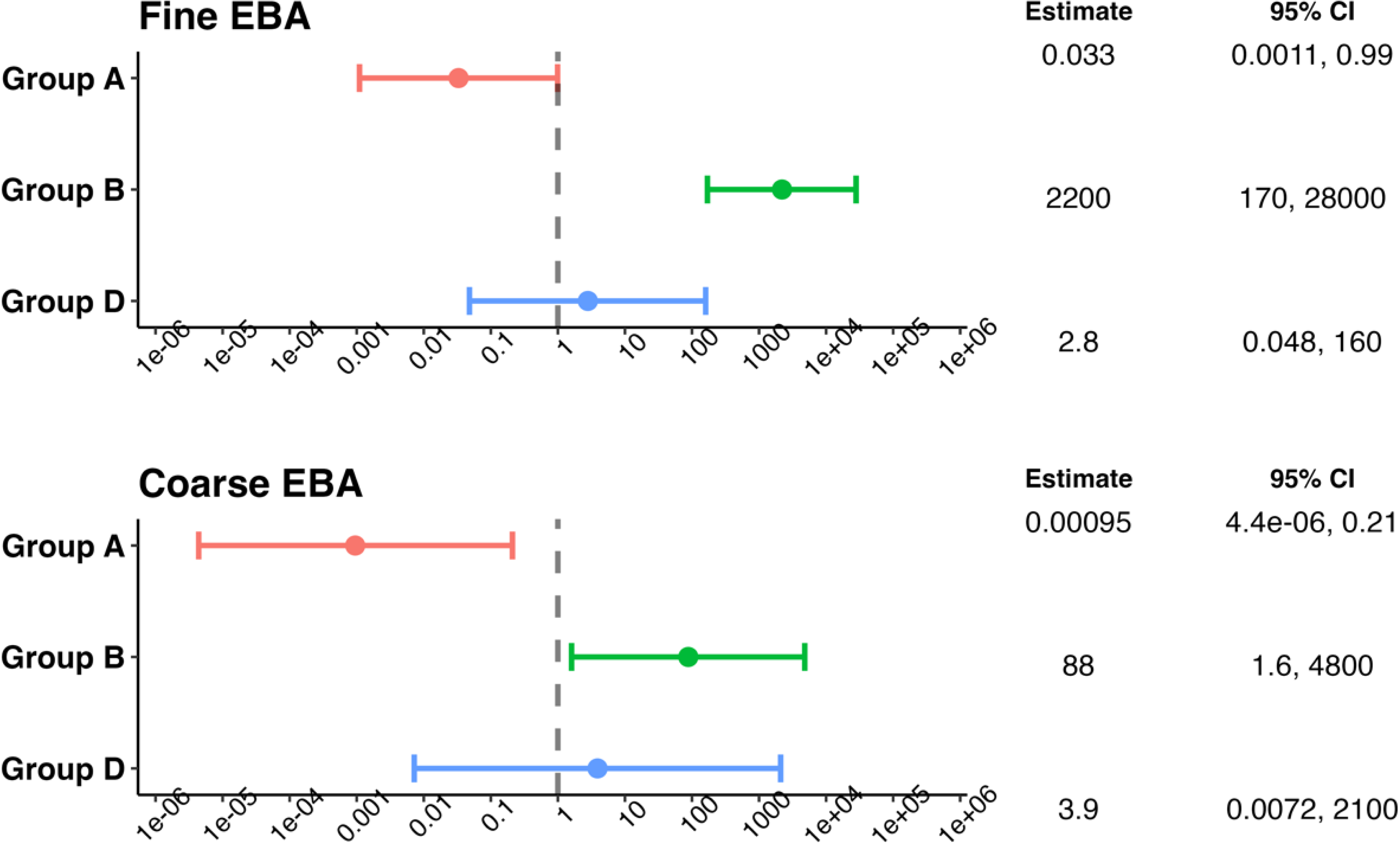
The ratio of viral shedding in EBA of selected and inoculated cases to shedding by unselected cases (Group B1), controlling for age, sex, and study days. Group A: Nasal Inoculation with influenza A/Wisconsin/67/2005; Group B: Symptomatic with T>37.8°C or positive antigen test; Group C: Surveillance cohort with any symptoms and positive PCR; Group D: Symptomatic, seeking medical care. We included only those samples whose study day was not missing (three C and one D cases were excluded due to not having symptom onset date on file or having no symptoms over the course of the follow-up period). We used linear mixed-effects models for censored responses (R Project package “lmec”) to estimate the ratio of viral shedding in EBA of Group A/B/D to Group C. These models accounted for the censored outcome variable as well as nested random effects of individuals and samples within the same individuals. The best models were selected based on Akaike Information Criterion (AIC).

Regarding shedding trajectories over time, no significant difference was observed between Group C and the rest of the groups concerning the dynamics of EBA viral RNA shedding by study days (Supplementary Table 4).

## Discussion

This study provides a unique comparison of EBA viral shedding pattern among influenza A/H3 infected individuals in distinct groups: nasally inoculated cases, unselected community-acquired cases from a surveillance cohort, and selected natural cases from different influenza seasons. We found previously that selected natural cases shed higher viral load in EBAs and were more symptomatic than experimental cases (8). This study further adds that unselected community-acquired cases, representing a wider spectrum of symptom severity, shed more viral RNA in their exhaled breath aerosols and were more symptomatic than the experimental, nasally inoculated cases as well, but on average shed less viral RNA than the selected natural cases that were more symptomatic and from different influenza season.

Those inoculated nasally with a high dose of influenza virus A/Wisconsin/67/2005 strain had a high percentage of people who did not have detectable level of viral RNA load in their exhaled breath aerosols or have any symptoms, despite shedding substantial virus into nasopharyngeal swabs. This indicates that nasal inoculation with this strain may not provide a robust model for understanding EBA viral shedding patterns or the range of symptom severity typical in natural influenza virus infections. Given the major role of aerosol inhalation in influenza transmission (6,7), inoculation via aerosol inhalation might offer a more representative model for mimicking natural influenza infections (13). Previous studies showed that inhalation of various strains of influenza viruses led to fevers in human subjects, a response rarely seen with intranasal instillation (14). Also, animal studies have demonstrated that aerosol inhalation of recombinant influenza viruses results in a more efficient infection of the lower respiratory tract and faster viral replication compared to intranasal inoculation (15). This might explain the lower viral load found in the exhaled breath aerosols of individuals inoculated intranasally, as opposed to those infected naturally.

Of note, despite sharing a similar characteristic and symptomatic profile, the two groups of medically attended cases from different years (Groups B and D) exhaled statistically different fine aerosol viral RNA loads. The discrepancy in the relationship between viral shedding and symptoms could be related to year-to-year differences in circulating strains of influenza A(H3). The elevated viral load in fine aerosols observed in participants from the 2012-2013 influenza season might, in part, explain the heavier influenza burden during that season compared to the 2018-2019 influenza season in the United States (5). This discovery underscores the importance of monitoring influenza virus strains, even within the same influenza A(H3) subtype, to better inform public health measures to mitigate the impact of future influenza seasons and protect vulnerable populations more effectively.

This study had several strengths. We did a unique comparison of EBA viral shedding pattern among influenza A(H3) infected individuals from three sources, allowing us to compare the viral shedding between experimental infections and the full range of ambulatory community-acquired influenza virus infections. Including participants from different years also allowed us to explore the difference between different strains of influenza A(H3).

Our research was subject to some limitations. Our study volunteers were all ambulatory; hence the results of this study may not be generalizable to those who are critically ill. We did not have culture results for all the EBA samples, hence we could not directly compare infectious viral load, and potentially the risk of inhalation transmission, among the groups. Despite this, previous work reported a correlation between viral RNA and quantitative culture (7).

This study reveals that the route of infection and strain difference could contribute to the level of viral aerosol shedding in influenza virus infections. We highlight the need for new models to study viral aerosol shedding from the human respiratory tract. In particular, aerosol inoculation may represent a promising avenue for future research, as this approach could better mimic natural infection process and offer insights into aerosol transmission dynamics. Overall, this study contributes to the broader understanding of viral shedding patterns in EBA and underscores the importance of developing robust models to further our understanding of influenza transmission.

## Materials and Methods

### Data sources and sample groups

This study used data obtained from three studies. The first two of these studies - Evaluating Modes of Influenza Transmission challenge study (EMIT-1 experimental study) (16), and an observational study of selected acute respiratory illness (ARI) cases (EMIT-1 campus study) (7) - have been previously reported (8). The third study followed uninfected volunteers over an academic year for their ARI occurrence, known as the Prometheus study (12).

### EMIT-1 experimental study

This study, initiated in 2013 to investigate questions related to influenza transmission (16), involved volunteers divided into two groups: “Donors” and “Recipients”. The Donors were intranasally inoculated with a dose of 5.5 log_10_TCID_50_ influenza A/Wisconsin/67/2005 and subsequently confirmed to have an influenza A infection (H3N2). After inoculation, the Donors were paired with uninfected healthy Recipients in a common room to encourage interaction (16). For the purposes of this report, we exclusively included the Donors, representing experimental cases, and have designated them as Group A.

### EMIT-1 campus study

The EMIT-1 campus study was conducted at the University of Maryland, College Park (UMD) (7,8). Volunteers presenting symptoms of acute respiratory infections were recruited from December 2012 to March 2013. Those who eventually provided breath samples were within 3 days of symptom onset, and had a positive QuickVue Influenza A + B test (Quidel, San Diego, CA) or were still febrile with an oral temperature of greater than 37.8 °C plus coughing or a sore throat (7,8). In the context of our current study, we incorporated those with a positive nasopharyngeal swab for influenza A/H3N2 as confirmed by reverse transcription polymerase chain reaction (RT-PCR). This group, representing the severe spectrum of symptoms, was classified as Group C.

### Prometheus study

The Prometheus study was also conducted at the UMD community (12). From 2017 to 2020, students and staff from UMD were actively recruited and monitored for the occurrence of acute respiratory infections during each academic year. Volunteers were instructed to report to the study team immediately upon developing cold or flu-like symptoms. The identified volunteers and their close contacts were then invited to a research clinic to provide samples for further examination. For the 2018-2019 flu season, additional volunteers were recruited from the university health center, typically presenting with more severe symptoms than those recruited from the surveillance cohort. For this study, we included individuals with confirmed influenza A via mid-turbinate or nasal and/or throat swabs, as identified by a TaqMan® Array Card (Thermo Fisher, Waltham, MA, USA). Participants from the community, generally presenting with very mild symptoms, were assigned to Group B1, while those recruited from the university health center were classified as Group B2.

### Sample Collection

For all these studies, volunteers with confirmed viral infections were invited to provide 30-minute EBA samples using a Gesundheit-II sampler (17). They were allowed to breathe normally and cough spontaneously during the collection. After collection, EBA samples were categorized by size into two fractions: fine aerosols (diameter ≤5 μm) and coarse aerosols (>5 μm). Viral RNA was extracted from both fine and coarse aerosol samples and quantified using real-time RT-PCR.

### Statistical analyses

In our statistical analysis, we included only EBA samples on which day their nasal and/or throat swabs were positive.

Descriptive analysis was carried out for the four comparison groups (A, B1, B2, and C). We compared the four groups and the three natural infection groups using Kruskal-Wallis test for the continuous variables and Fisher’s exact test for the categorical variables. We also did pair-wise comparison among the three natural infection groups, using T-test for the continuous variables and Fisher’s exact test or Chi-square test for the categorical variables.

To visualize change over time, we plotted the EBA viral RNA loads as well as symptom scores by study day for the four groups separately. The study day was defined as days post symptom onset for Group B1, B2, and C and days post inoculation for Group A, as most of the experimental cases were asymptomatic or not sampled on the days after symptom onset. We then identified the incubation period (*I*) for Group A by comparing the peak of symptom scores in Group A and the other three groups.

Linear mixed effect models with censored responses (R package ‘lmec’, version 1.0 (18)) were used to calculate the geometric means (GM) of the EBA viral RNA load for the four comparison groups. These models accounted for the censored outcome variable (i.e., viral RNA load below limit of detection) as well as nested random effects of individuals and samples within the same individuals. We also used the lmec model to estimate the group effect on EBA viral shedding (Model I) and the shedding over time (Model II), controlling for potential confounders and effect modifiers. For Model I, age, sex, and study day (defined as day post symptom onset for Group B1, B2, and C, and day post inoculation minus *I* for Group C) were included into the models as potential confounders. Then the product terms of each of these three variables and the group variable were included for further interaction assessment. The best models were selected based on Akaike Information Criterion (AIC). For Model II, we forced the interaction terms of group and study day into the model along with age and sex, and then further selected the final model based on AIC.

We conducted all analyses in R version 4.2.3 (R Foundation for Statistical Computing, Vienna, Austria) and RStudio.

## Data Availability

Data will be made available in a publicly accessible repository upon acceptance of the manuscript.

## Acknowledgement

We thank all the members of the EMIT Consortium and Prometheus-UMD investigators (Both complete lists of the members can be found in the Supporting Materials).

## Supplementary materials

### EMIT Consortium Team Members

EMIT team members were: Walt Adamson, Blanca Beato-Arribas, Werner Bischoff, William Booth, Simon Cauchemez, Sheryl Ehrman, Joanne Enstone, Neil Ferguson, John Forni, Anthony Gilbert, Michael Grantham, Lisa Grohskopf, Andrew Hayward, Michael Hewitt, Ashley Kang, Ben Killingley, Robert Lambkin-Williams, Alex Mann, Donald Milton, Jonathan Nguyen-Van-Tam, Catherine Noakes, John Oxford, Massimo Palmarini, Jovan Pantelic, and Jennifer Wang. The Scientific Advisory Board members were: Allan Bennett, Ben Cowling, Arnold Monto, and Raymond Tellier.

### Prometheus-UMD investigators

Addo, Kofi

Adenaiye, Oluwasanmi Oladapo

Agrawala, Agrawala

Aiello, Allison

Albert, Barbara

Arria, Amelia

Bueno de Mesquita, P. Jacob

Cai, Mara

Chen, Shuo

Chen, Wilbur

Corrada Bravo, Hector

Elworth, Leo

Felgner, Philip

Frieman, Matthew

German, Jennifer

Heidarinejad, Mohammad

Hong, Filbert

Jiang, Chengsheng

Khan, Saahir

Lai, Jianyu

Liu, Hongjie

Ma, Tianzhou

Maljkovic Berry, Irina

Martinello, Richard

Mattise, Nick

Memon, Atif

Milton, Donald

Mongodin, Emmanuel

Nasko, Dan

Pop, Mihai

Porter, Adam

Romo, Sebastian

Srebric, Jelena

Tai, Sheldon

Treangen, Todd

Wajid, Faizan

Washington-Lewis, Rhonda

Wu, Qiong

Xing, Yishi

Youssefi, Somayeh

Zhu, Shengwei

### Supplementary Tables

**Supplementary Table 1.** Group comparisons for the Characteristics of the study population.

|  | 4-group<br>comparison<sup>a</sup><br>(A, B, C, D),<br>*p* | 3-group<br>comparison<br>(B, C, D),<br>*p* | C vs. D<sup>b</sup>,<br>*p* | B vs. C,<br>*p* | B vs. D,<br>*p* |
| --- | --- | --- | --- | --- | --- |
| Flu season | <0.001 | <0.001 | 0.222 | <0.001 | <0.001 |
| Female | 0.02 | 0.293 | 0.182 | 0.73 | 0.239 |
| Age | <0.001 | 0.008 | 0.159 | <0.001 | 0.433 |
| Age group | <0.001 | 0.695 | 1 | 0.549 | 1 |
| With fever ><br>37.9°C | 0.056 | 0.896 | 1 | 0.73 | 1 |
| Coughs per 30<br>min | <0.001 | 0.042 | 0.826 | 0.08 | 0.338 |
| Temperature<br>(C) | <0.001 | 0.14 | 0.32 | 0.711 | 0.256 |
| Upper<br>respiratory<br>symptoms | <0.001 | 0.061 | 0.87 | 0.028 | 0.139 |
| Lower<br>respiratory<br>symptoms | <0.001 | 0.403 | 0.498 | 0.146 | 0.939 |
| Systemic<br>symptoms | <0.001 | 0.004 | 0.08 | <0.001 | 0.957 |

- 547 a. Four-group and three-group comparison: Kruskal-Wallis test for the continuous variables and Fisher's exact test for the categorical variables - 549 b. Two-group comparison: T-test for the continuous variables and Fisher's exact test or Chi-square test for the categorical variables.

**Supplementary Table 2.** The effect of group (ref = Group C) on EBA viral RNA shedding^a,b^.

| Variables |  | Fine EBA | Coarse EBA |
| --- | --- | --- | --- |
| Group<sup>b</sup>(Ref=C) | A | 0.033 (0.0011, 0.99) | 0.00095 (4.4e-06, 0.21) |
|  | B | 2200 (170, 28000) | 88 (1.6, 4800) |
|  | D | 2.8 (0.048, 160) | 3.9 (0.0072, 2100) |
| Study Day<sup>c</sup><br>(DPS/DPI-2) |  | 0.27 (0.14, 0.52) | 0.19 (0.079, 0.46) |
| Age |  | 1.1 (0.95, 1.2) | 1.1 (0.94, 1.3) |
| Sex |  | 8.2 (1.8, 36) | 6.4 (0.65, 62) |

**Supplementary Table 3.**
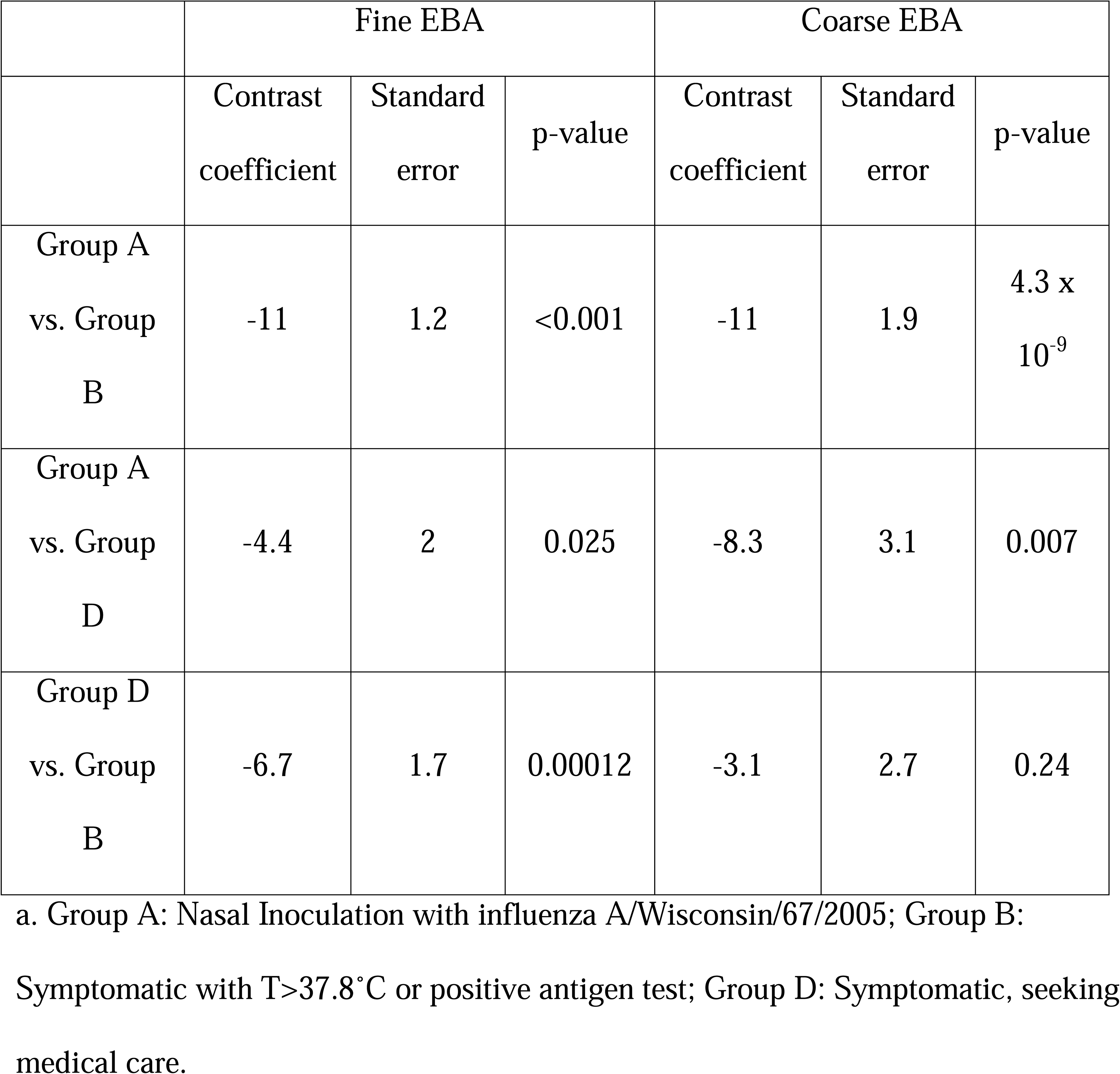
Contrast analysis on the relative effect of selected and inoculated cases^a^.

|  | Fine EBA |  |  | Coarse EBA |  |  |
| --- | --- | --- | --- | --- | --- | --- |
|  | Contrast<br>coefficient | Standard<br>error | p-value | Contrast<br>coefficient | Standard<br>error | p-value |
| Group A<br>vs. Group<br>B | -11 | 1.2 | <0.001 | -11 | 1.9 | 4.3 x<br>10<sup>-9</sup> |
| Group A<br>vs. Group<br>D | -4.4 | 2 | 0.025 | -8.3 | 3.1 | 0.007 |
| Group D<br>vs. Group<br>B | -6.7 | 1.7 | 0.00012 | -3.1 | 2.7 | 0.24 |

Symptomatic with T>37.8°C or positive antigen test; Group D: Symptomatic, seeking
medical care.

**Supplementary Table 4.** The effect of group (ref = Group C) on EBA viral RNA shedding over study day^a,b,c^.

| Variables |  | Fine EBA | Coarse EBA |
| --- | --- | --- | --- |
| Group<br>(Ref=C) | A | 0.0052 (1.7e-05, 1.6) | 3.7 (4.6e-05, 310000) |
|  | B | 1200 (4.9, 280000) | 720000 (13, 4e+10) |
|  | D | 0.25 (4.5e-06, 14000) | 11000 (0.00017, 6.5e+11) |
| Study Day (DPS/DPI-2) |  | 0.13 (0.0098, 1.8) | 10 (0.11, 910) |
| Age |  | 1.1 (0.95, 1.2) | 1.1 (0.96, 1.3) |
| Sex |  | 8.5 (1.9, 38) | 4.7 (0.49, 45) |
| Group A x Study Day |  | 10 (0.51, 210) | 0.019 (0.00011, 3.4) |
| Group B x Study Day |  | 1.5 (0.096, 22) | 0.012 (0.00012, 1.2) |
| Group D x Study Day |  | 4.7 (0.0035, 6300) | 0.027 (3.7e-07, 1900) |

### Supplementary Figure

**Supplementary Figure 1.**
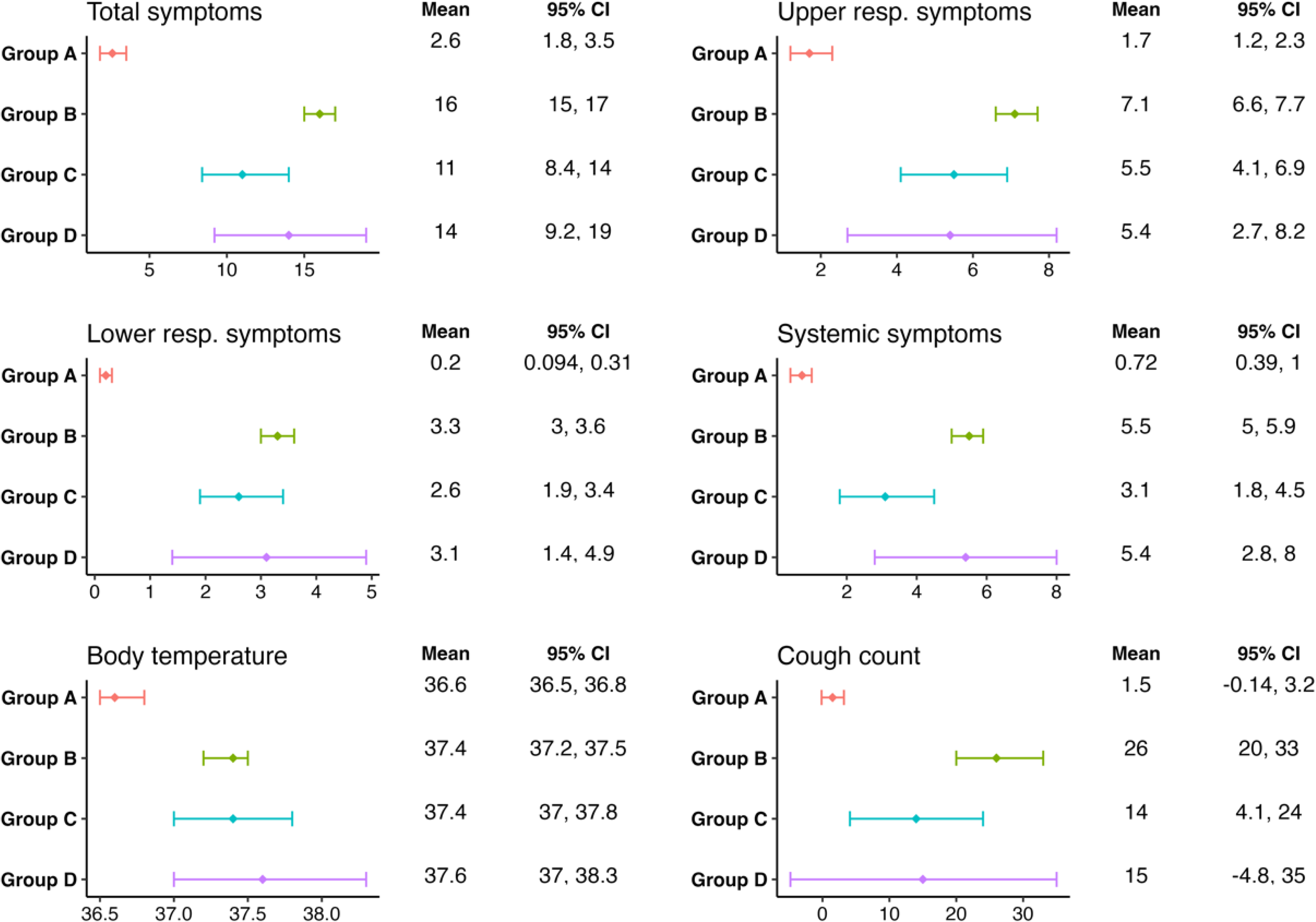
Mean symptom scores, body temperatures, and cough counts across groups. Group A: Nasal Inoculation with influenza A/Wisconsin/67/2005; Group B: Symptomatic with T>37.8°C or positive antigen test; Group C: Surveillance cohort with any symptoms and positive PCR; Group D: Symptomatic, seeking medical care.

